# Sociodemographic and Household Environmental Determinants of Stunting at Birth: Evidence of Regional Disparities in Indonesia

**DOI:** 10.64898/2026.07.01.26357022

**Authors:** Fawwiz Aulya Amin, Siti Helmyati, Vicka Oktaria

## Abstract

**Background:** Stunting at birth reflects impaired fetal growth and increases the risk of adverse health outcomes throughout the life course. Evidence on regional variations in determinants of stunting at birth in Indonesia remains limited. This study identified sociodemographic, and household environmental determinants of stunting at birth at both the national and regional levels.

**Methods:** This cross-sectional study analyzed data from the 2024 Indonesian Nutritional Status Survey (SSGI), including 28,982 children aged 0–23 months. Bivariate and multivariable logistic regression analyses were conducted nationally and across seven regions: Sumatra, Java–Bali, Nusa Tenggara, Kalimantan, Sulawesi, Maluku, and Papua. Adjusted odds ratios (aORs) and 95% confidence intervals (CIs) were reported.

**Results:** The prevalence of stunting at birth was 5.53%, with the highest prevalence in Sulawesi (8.02%). Nationally, household crowding was the only factor significantly associated with stunting at birth after adjustment. Regional analyses revealed considerable variation in associated factors. In Sumatra, urban residence was associated with lower odds of stunting at birth (aOR: 0.73; 95% CI: 0.55–0.97), while children from the lowest wealth quintile had higher odds (aOR: 1.78; 95% CI: 1.03–3.06). In Java–Bali, low maternal nutritional knowledge was associated with increased odds of stunting at birth (aOR: 1.65; 95% CI: 1.02–2.66). In Nusa Tenggara, limited drinking water services (aOR: 2.35; 95% CI: 1.09–5.07) and limited sanitation services (aOR: 2.34; 95% CI: 1.28–4.26) were associated with higher odds of stunting at birth.

**Conclusions:** Determinants of stunting at birth differed across Indonesian regions. While household crowding emerged as a national determinant, socioeconomic factors were more important in Sumatra, maternal nutritional knowledge in Java–Bali, and environmental conditions in Nusa Tenggara. These findings highlight the need for region-specific strategies to prevent stunting from the prenatal period.

## Introduction

Stunting remains a major public health challenge worldwide, affecting an estimated 148 million children under five years of age in 2022. Despite global efforts to reduce its prevalence, progress remains insufficient, particularly in low- and middle-income countries. In Indonesia, stunting has become a national development priority. Although the prevalence of stunting declined from 21.5% in 2023 to 19.8% in 2024, it remains above the national target of 14%. Evidence also suggests that growth faltering may begin before birth. Using data from the 2018 Basic Health Research Survey, Sari and Sartika (2021) reported that 10.2% of Indonesian newborns were already stunted at birth. In addition, the proportion of infants born with a birth length below 48 cm remained high, decreasing only slightly from 19.4% in 2021 to 18.4% in 2024 [2], [3].

Stunting at birth is defined as a birth length-for-age Z-score below -2 standard deviations according to the WHO Child Growth Standards [4]. As an indicator of impaired fetal growth, stunting at birth reflects adverse conditions experienced during pregnancy and may serve as an early predictor of long-term nutritional status. Infants born stunted face a higher risk of persistent growth failure, impaired cognitive development, reduced educational attainment, and lower productivity later in life [5]. Furthermore, stunted newborns are four times more likely to remain stunted at three months of age and twice as likely to be stunted at two years of age. They also tend to experience limited catch-up growth, resulting in shorter adult stature [6].

In addition to biological determinants, sociodemographic and household environmental factors may influence fetal growth and birth outcomes. Previous studies have shown that parental education, health-related knowledge, household economic status, and place of residence affect access to health information, healthcare services, and nutritional resources. Likewise, household environmental conditions, including access to safe drinking water, sanitation facilities, and adequate housing, may influence health through increased exposure to infectious diseases and poor living conditions [7], [8], [9], [10], [11]. However, most existing studies have focused on stunting among children under five, while evidence regarding the contribution of sociodemographic and household environmental factors to stunting at birth remains limited.

Indonesia is characterized by substantial geographic, socioeconomic, and environmental diversity across regions. These differences may contribute to regional variations in the determinants of stunting at birth. Nevertheless, evidence examining regional disparities in sociodemographic and household environmental determinants of stunting at birth remains scarce. Most studies have focused on national-level estimates, which may mask important contextual differences across regions.

Therefore, this study aimed to examine the association between sociodemographic and household environmental factors and stunting at birth using data from the 2024 Indonesian Nutritional Status Survey (SSGI). In addition, this study explored regional disparities in these associations to provide evidence for more targeted and context-specific strategies to prevent stunting from the earliest stages of life.

## Method

### Study Design and Data Source

This study employed a cross-sectional design using secondary data from the 2024 Indonesian Nutritional Status Survey (Survei Status Gizi Indonesia/SSGI). The SSGI is a nationally representative survey conducted by the Ministry of Health of Indonesia to monitor nutritional status and determinants of maternal and child health across all provinces.

### Study Population and Sample

The study population consisted of children aged 0–59 months included in the 2024 SSGI. Children with complete information on birth length and study variables were eligible for inclusion in the analysis. Children with missing birth length data, preterm birth, or incomplete information on study variables were excluded from the analysis. A total of 28,982 children met the inclusion criteria and were included in the final analysis.

**Figure 1.**
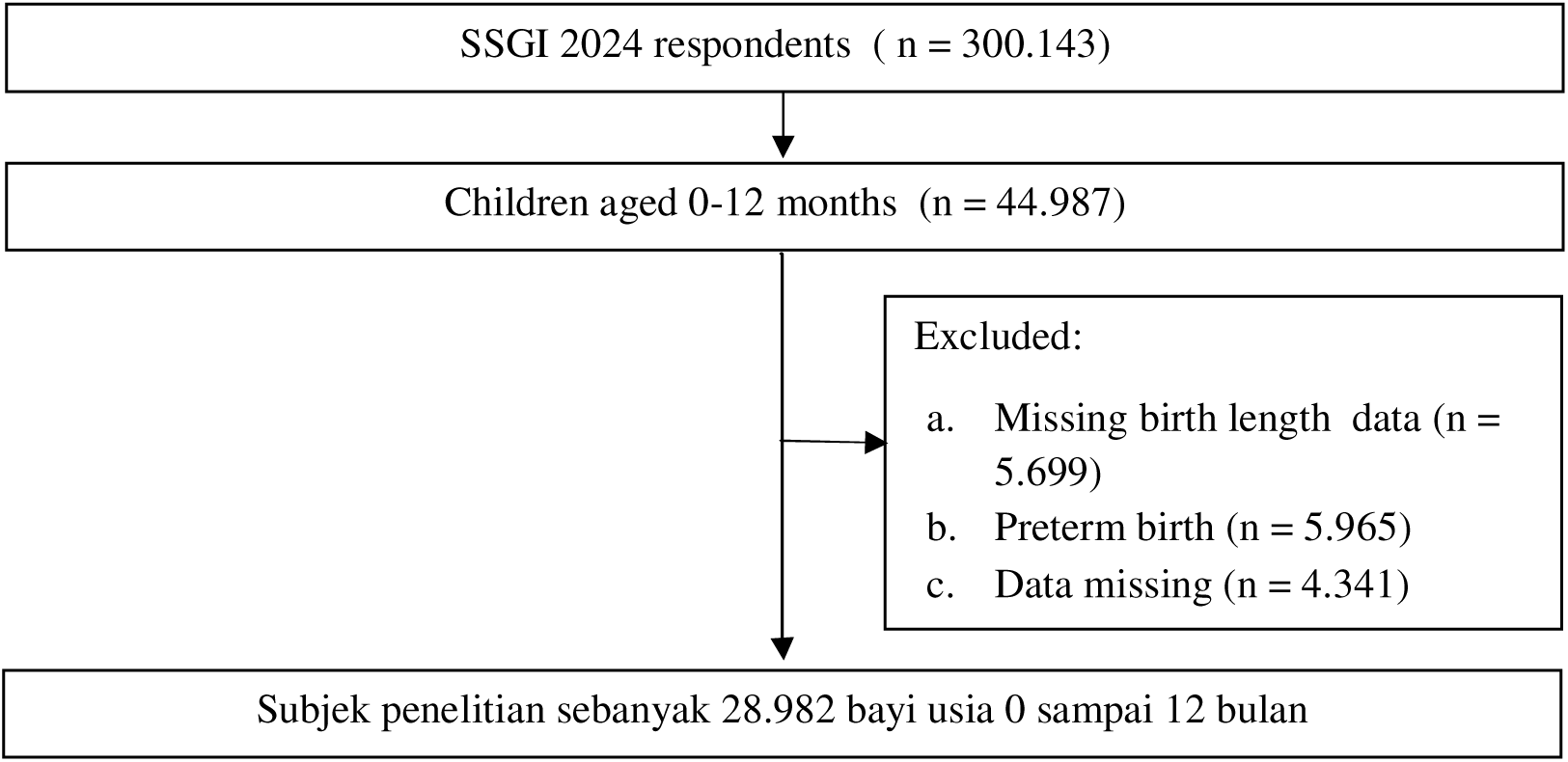
Flowchart of participant selection from the 2024 Indonesian Nutritional Status Survey (SSGI).

### Variables

The dependent variable was stunting at birth. Stunting at birth was defined as birth length-for-age below -2 standard deviations according to the WHO Child Growth Standards [4]. The independent variables comprised sociodemographic and household environmental factors. Sociodemographic factors included maternal education, paternal education, maternal knowledge regarding stunting, household economic status, and place of residence. Household environmental factors included drinking water access, sanitation access, and household crowding.

Maternal education and paternal education were categorized into low and high education levels. Maternal knowledge regarding stunting was classified as low or high based on the survey scoring system. Household economic status was categorized into five wealth quintiles. Place of residence was classified as urban or rural. Drinking water access and sanitation access were categorized according to the WHO/UNICEF Joint Monitoring Programme (JMP) classifications, while household crowding was categorized as crowded and non-crowded based on national housing standards.

### Statistical Analysis

Descriptive statistics were used to summarize respondent characteristics and estimate the prevalence of stunting at birth. Bivariable associations between each independent variable and stunting at birth were examined using survey logistic regression. Variables with p-values <0.25 in the bivariable analysis and variables considered theoretically relevant were included in the multivariable analysis. Multivariable survey logistic regression was performed to estimate adjusted odds ratios (AORs) and 95% confidence intervals (CIs) for sociodemographic and household environmental factors associated with stunting at birth. To assess regional disparities, separate multivariable analyses were conducted for seven Indonesian regions: Sumatra, Java–Bali, Kalimantan, Sulawesi, Nusa Tenggara, Maluku, and Papua. All analyses accounted for the complex survey design and sampling weights. Statistical significance was determined at p <0.05. Several measures were undertaken to minimize potential sources of bias. Selection bias was reduced by using data from a nationally representative survey with standardized sampling procedures. Information bias was minimized through the use of standardized questionnaires and measurement protocols implemented by trained field enumerators.

### Ethical Considerations

Ethical approval was obtained from the Medical and Health Research Ethics Committee, Faculty of Medicine, Public Health and Nursing, Universitas Gadjah Mada, Indonesia (No. KE/FK/0357/EC/2026; 24 February 2026). This study used anonymized secondary data from the 2024 Indonesian Nutritional Status Survey (SSGI), and participant confidentiality was maintained throughout the study.

## Results

A total of 28,982 children aged 0–23 months were included in the analysis. The overall prevalence of stunting at birth was 5.53%. Table 1 presents the distribution of ociodemographic, household, and environmental characteristics of the study population.

**Table 1.**
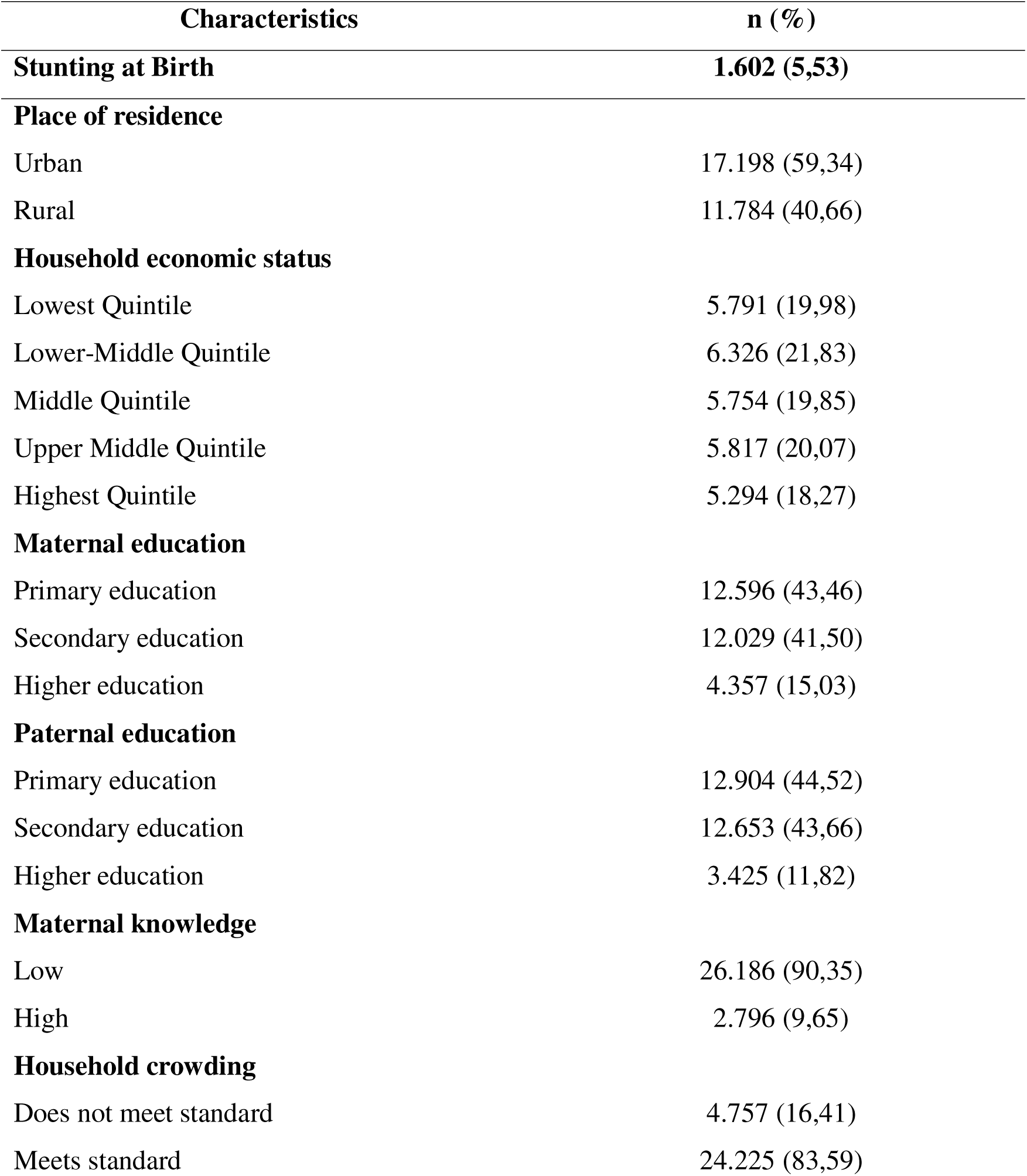
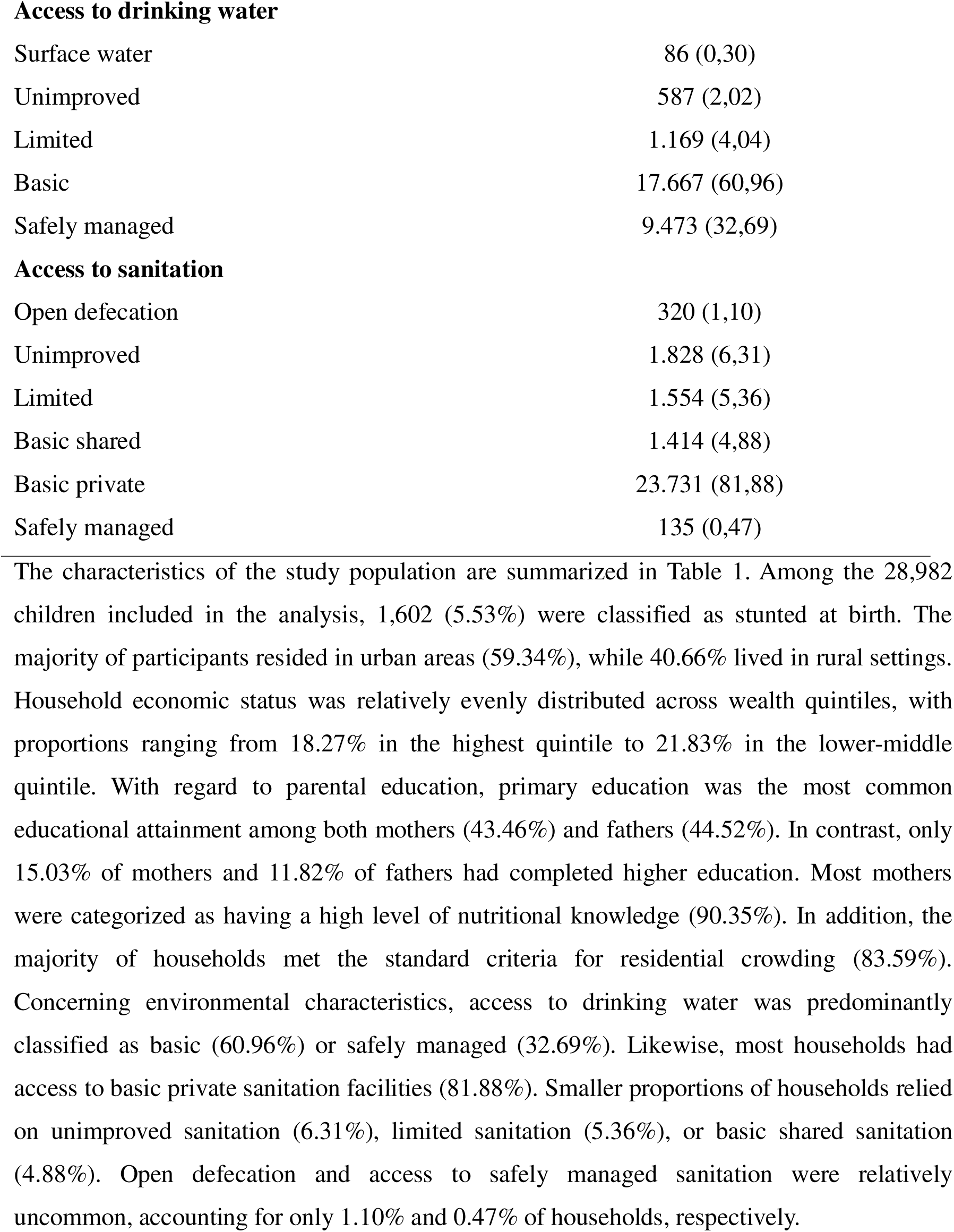
Sociodemographic, Household, and Environmental Characteristics of the Study Population, Indonesia, 2022 (N=28,982)

| Characteristics | n (%) |
| --- | --- |
| **Stunting at Birth** | **1.602 (5,53)** |
| **Place of residence** |  |
| Urban | 17.198 (59,34) |
| Rural | 11.784 (40,66) |
| **Household economic status** |  |
| Lowest Quintile | 5.791 (19,98) |
| Lower-Middle Quintile | 6.326 (21,83) |
| Middle Quintile | 5.754 (19,85) |
| Upper Middle Quintile | 5.817 (20,07) |
| Highest Quintile | 5.294 (18,27) |
| **Maternal education** |  |
| Primary education | 12.596 (43,46) |
| Secondary education | 12.029 (41,50) |
| Higher education | 4.357 (15,03) |
| **Paternal education** |  |
| Primary education | 12.904 (44,52) |
| Secondary education | 12.653 (43,66) |
| Higher education | 3.425 (11,82) |
| **Maternal knowledge** |  |
| Low | 26.186 (90,35) |
| High | 2.796 (9,65) |
| **Household crowding** |  |
| Does not meet standard | 4.757 (16,41) |
| Meets standard | 24.225 (83,59) |
| **Access to drinking water** |  |
| Surface water | 86 (0,30) |
| Unimproved | 587 (2,02) |
| Limited | 1.169 (4,04) |
| Basic | 17.667 (60,96) |
| Safely managed | 9.473 (32,69) |
| **Access to sanitation** |  |
| Open defecation | 320 (1,10) |
| Unimproved | 1.828 (6,31) |
| Limited | 1.554 (5,36) |
| Basic shared | 1.414 (4,88) |
| Basic private | 23.731 (81,88) |
| Safely managed | 135 (0,47) |

**Table 2.** Association of Sociodemographic, Household, and Environmental Factors with Stunting at Birth in Indonesia

| Variables | Stunting at birth (%) | cOR (95% CI) | p-value | aOR (95% CI) | p-value |
| --- | --- | --- | --- | --- | --- |
| **Place of residence** |  |  |  |  |  |
| Urban | 3,40 | 0,90 (0,77-1,05) | 0,212 | 0,91 (0,78-1,07) | 0,282 |
| Rural | 2,12 | 1,00 (reference) |  | 1,00 (reference) |  |
| **Household economic status** |  |  |  |  |  |
| Lowest Quintile | 1,19 | 1,16 (0,91-1,48) | 0,207 | 1,21 (0,91-1,59) | 0,174 |
| Lower-Middle Quintile | 1,16 | 1,03 (0,82-1,31) | 0,755 | 1,08 (0,83-1,41) | 0,524 |
| Middle Quintile | 1,08 | 1,05 (0,83-1,35) | 0,643 | 1,11 (0,86-1,45) | 0,397 |
| Upper Middle Quintile | 1,15 | 1,12 (0,86-1,46) | 0,34 | 1,17 (0,90-1,53) | 0,231 |
| Highest Quintile | 0,94 | 1,00 (reference) |  | 1,00 (reference) |  |
| **Maternal education** |  |  |  |  |  |
| Primary education | 2,40 | 0,96 (0,78-1,19) | 0,775 | 0,94 (0,72-1,23) | 0,681 |
| Secondary education | 2,27 | 0,95 (0,77-1,18) | 0,689 | 0,95 (0,75-1,20) | 0,694 |
| Higher education | 0,86 | 1,00 (reference) |  | 1,00 (reference) |  |
| **Paternal education** |  |  |  |  |  |
| Primary education | 2,28 | 0,93 (0,73-1,18) | 0,564 | 0,89 (0,65-1,22) | 0,484 |
| Secondary education | 2,35 | 0,89 (0,70-1,14) | 0,379 | 0,86 (0,66-1,12) | 0,288 |
| Higher education | 0,70 | 1,00 (reference) |  | 1,00 (reference) |  |
| **Maternal knowledge** |  |  |  |  |  |
| Low | 5,05 | 1,15 (0,90-1,46) | 0,242 | 1,15 (0,90-1,46) | 0,243 |
| High | 0,47 | 1,00 (reference) |  | 1,00 (reference) |  |
| **Household crowding** |  |  |  |  |  |
| Does not meet standard | 4,42 | 1,29 (1,06-1,56) | 0,008 | 1,27 (1,04-1,54) | 0,017 |
| Meets standard | 1,10 | 1,00 (reference) |  | 1,00 (reference) |  |
| **Access to drinking water** |  |  |  |  |  |
| Surface water | 0,02 | 1,01 (0,40-2,50) | 0,981 | 0,97 (0,39-2,43) | 0,963 |
| Unimproved | 0,10 | 0,87 (0,54-1,39) | 0,570 | 0,86 (0,53-1,38) | 0,537 |
| Limited | 0,24 | 1,07 (0,76-1,51) | 0,660 | 1,06 (0,75-1,49) | 0,725 |
| Basic | 3,39 | 1,02 (0,86-1,20) | 0,787 | 1,00 (0,85-1,18) | 0,929 |
| Safety managed | 1,78 | 1,00 (reference) |  | 1,00 (reference) |  |
| **Access to sanitation** |  |  |  |  |  |
| Open defecation | 0,04 | 0,75 (0,31-1,65) | 0,445 | 0,69 (0,30-1,59) | 0,393 |
| Unimproved | 0,39 | 1,14 (0,55-2,33) | 0,719 | 1,13 (0,55-2,32) | 0,737 |
| Limited | 0,35 | 1,19 (0,59-2,42) | 0,618 | 1,15 (0,57-2,34) | 0,684 |
| Basic shared | 0,26 | 0,97 (0,46-2,01) | 0,941 | 0,95 (0,46-1,98) | 0,909 |
| Basic private | 4,45 | 0,98 (0,51-1,89) | 0,975 | 1,01 (0,52-1,94) | 0,964 |
| Safely managed | 0,03 | 1,00 (reference) |  | 1,00 (reference) |  |

The distribution of study participants across seven regions in Indonesia is presented in Table 3. More than half of the participants were located in the Java–Bali region (57.15%), followed by Sumatra (21.79%). The overall prevalence of stunting at birth was 5.53%, with the highest prevalence observed in Sulawesi (8.02%). Regional differences were also evident in place of residence. Urban residence was more prevalent in Java–Bali (66.93%) and Papua (64.13%), whereas rural residence predominated in Sumatra, Nusa Tenggara, Sulawesi, and Maluku. Household economic status showed a relatively even distribution across most regions; however, a substantial proportion of households in Nusa Tenggara fell within the lowest wealth quintile (46.04%). Maternal and paternal education levels were generally concentrated at the primary and secondary education levels across all regions. The majority of households met the standard for residential crowding (83.59%), although Nusa Tenggara recorded the highest proportion of overcrowded households (31.51%). Access to drinking water was largely categorized as basic (60.96%) or safely managed (32.69%). Similarly, most households had access to basic private sanitation facilities (81.88%), while open defecation remained uncommon (1.10%). However, marked regional disparities were observed. Maluku and Papua exhibited the highest proportions of households relying on limited drinking water services (18.37% and 11,67, respectively), while access to safely managed drinking water was substantially lower in Sulawesi (24.05%) and Papua (24.33%) than in other regions. In terms of sanitation, Nusa Tenggara had the highest proportion of households using limited sanitation facilities (15.88%) and the lowest coverage of basic private sanitation (75.82%), whereas unimproved sanitation was most prevalent in Java–Bali (8.21%) and Kalimantan (6.17%).

**Table 3.** Sociodemographic, Household, and Environmental Characteristics of Study Participants Across Regions in Indonesia

| Variabel | Sumatera | Jawa-Bali | Nusa<br>Tenggara | Kalimantan | Sulawesi | Maluku | Papua | Total |
| --- | --- | --- | --- | --- | --- | --- | --- | --- |
|  | n (%) | n (%) | n (%) | n (%) | n (%) | n (%) | n (%) | n (%) |
|  | 6.314 (21,79) | 16.563 (57,15) | 1,229 (4,24) | 2.100 (7,25) | 2.237 (7,72) | 190 (0,66) | 349 (1,20) | 28.982 (100) |
| **Stunting at birth** | 358 (5,68) | 848 (5,12) | 68 (5,57) | 118 (5,60) | 179 (8,02) | 10 (5,53) | 20 (5,61) | 1.601 (5,53) |
| **Place of residence** |  |  |  |  |  |  |  |  |
| Urban | 3.098 (49,06) | 11.085 (66,93) | 555 (45,19) | 1.113 (52,98) | 1.048 (46,86) | 75 (39,68) | 224 (64,13) | 17.198 (59,34) |
| Rural | 3.216 (50,94) | 5.478 (33,07) | 674 (54,81) | 987 (47,02) | 1.189 (53,14) | 115 (60,32) | 125 (35,87) | 11.784 (40,66) |
| **Household economic status** |  |  |  |  |  |  |  |  |
| Lowest Quintile | 1.055 (16,72) | 3.272 (19,75) | 566 (46,04) | 292 (13,91) | 441 (19,74) | 67 (35,16) | 98 (28,09) | 5.791 (19,98) |
| Lower-Middle Quintile | 1.192 (18,87) | 3.956 (23,88) | 252 (20,48) | 387 (18,43) | 465 (20,80) | 35 (18,61) | 39 (11,14) | 6.326 (21,83) |
| Middle Quintile | 1.436 (22,75) | 3.153 (19,04) | 169 (13,77) | 452 (21,51) | 438 (19,57) | 42 (22,03) | 64 (18,30) | 5.754 (19,85) |
| Upper Middle Quintile | 1.454 (23,03) | 3.192 (19,27) | 152 (12,37) | 479 (22,81) | 441 (22,03) | 24 (12,63) | 75 (21,39) | 5.817 (20,07) |
| Highest Quintile | 1.177 (18,63) | 2.990 (18,06) | 90 (7,34) | 490 (23,34) | 452 (18,30) | 22 (11,57) | 73 (21,08) | 5.294 (18,27) |
| **Maternal education** |  |  |  |  |  |  |  |  |
| Primary education | 2.373 (37,58) | 7.624 (46,03) | 541 (44,07) | 1.022 (48,64) | 880 (39,32) | 44 (23,24) | 112 (32,28) | 12.596 (43,46) |
| Secondary education | 2.927 (46,36) | 6.704 (40,48) | 437 (35,57) | 806 (38,39) | 875 (39,10) | 109 (57,19) | 171 (48,98) | 12.029 (41,50) |
| Higher education | 1.014 (16,06) | 2.235 (13,49) | 250 (20,36) | 273 (12,98) | 483 (21,58) | 37 (19,57) | 65 (18,74) | 4.357 (15,03) |
| **Paternal education** |  |  |  |  |  |  |  |  |
| Primary education | 2.619 (41,48) | 7.649 (46,18) | 562 (45,77) | 968 (46,06) | 951 (42,49) | 56 (29,40) | 99 (28,42) | 12.904 (44,52) |
| Secondary education | 2.996 (47,45) | 7.035 (42,48) | 450 (36,60) | 914 (43,51) | 962 (43,00) | 106 (56,04) | 190 (54,43) | 12.653 (43,66) |
| Higher education | 699 (11,07) | 1.878 (11,34) | 216 (17,63) | 219 (10,43) | 325 (14,52) | 28 (14,56) | 60 (17,15) | 3.425 (11,82) |
|  | n (%) | n (%) | n (%) | n (%) | n (%) | n (%) | n (%) | n (%) |
|  | 6.314 (21,79) | 16.563 (57,15) | 1,229 (4,24) | 2.100 (7,25) | 2.237 (7,72) | 190 (0,66) | 349 (1,20) | 28.982 (100) |
| **Maternal knowledge** |  |  |  |  |  |  |  |  |
| Low | 5.662 (89,67) | 15.153 (91,49) | 1.046 (85,15) | 1.851 (88,14) | 1.990 (88,95) | 164 (86,11) | 319 (91,63) | 26.185 (90,35) |
| High | 652 (10,33) | 1.410 (8,51) | 182 (14,85) | 249 (11,86) | 247 (11,05) | 27 (13,89) | 29 (8,37) | 2.796 (9,65) |
| **Household crowding** |  |  |  |  |  |  |  |  |
| Does not meet standard | 997 (15,79) | 2.551 (15,40) | 387 (31,51) | 302 (14,37) | 426 (19,07) | 18 (78,02) | 76 (21,98) | 4.757 (16,41) |
| Meets standard | 5.317 (84,21) | 14.012 (84,60) | 841 (68,49) | 1.799 (85,63) | 1.811 (80,93) | 173 (90,73) | 272 (78,02) | 24.225 (83,59) |
| **Access to drinking water** |  |  |  |  |  |  |  |  |
| Surface water | 15 (0,23) | 14 (0,08) | 7 (0,56) | 40 (1,91) | 3 (0,14) | 1 (0,42) | 6 (1,64) | 86 (0,30) |
| Unimproved | 138 (2,19) | 280 (1,69) | 54 (4,41) | 44 (2,09) | 59 (2,62) | 7 (3,57) | 5 (1,52) | 587 (2,02) |
| Limited | 330 (5,22) | 351 (2,12) | 88 (7,20) | 119 (5,68) | 206 (9,20) | 35 (18,37) | 41 (11,67) | 1.170 (4,04) |
| Basic | 3.395 (53,77) | 10.716 (64,70) | 517 (42,08) | 1.314 (62,55) | 1.432 (63,99) | 81 (42,79) | 212 (60,85) | 17.667 (60,96) |
| Safety managed | 2.436 (38,59) | 5.202 (31,41) | 562 (45,76) | 583 (27,77) | 538 (24,05) | 66 (34,85) | 85 (24,33) | 9.472 (32,69) |
| **Access to sanitation** |  |  |  |  |  |  |  |  |
| Open defecation | 103 (1,63) | 151 (0,91) | 15 (1,20) | 13 (0,63) | 32 (1,42) | 2 (0,83) | 4 (1,16) | 320 (1,10) |
| Unimproved | 282 (4,46) | 1.360 (8,21) | 26 (2,14) | 129 (6,17) | 20 (0,88) | 1 (0,35) | 10 (2,85) | 1.828 (6,31) |
| Limited | 213 (3,37) | 848 (5,12) | 195 (15,88) | 102 (4,88) | 168 (7,51) | 10 (5,24) | 18 (5,24) | 1.554 (5,36) |
| Basic shared | 183 (2,90) | 980 (5,91) | 54 (4,40) | 67 (3,19) | 96 (4,31) | 11 (6,47) | 23 (6,47) | 1.414 (4,88) |
| Basic private | 5.521 (87,44) | 13.150 (79,39) | 932 (75,82) | 1.748 (83,23) | 1.919 (85,80) | 167 (84,29) | 294 (84,29) | 23.731 (81,88) |
| Safely managed | 12 (0,19) | 74 (0,45) | 7 (0,56) | 40 (0,19) | 2 (0,08) | 0 | 0 | 135 (0,47) |

The crude associations between household characteristics and stunting at birth across seven regions of Indonesia are presented in Table 4. Overall, only a limited number of household characteristics demonstrated significant associations with stunting at birth, and the observed relationships varied across regions. Regional differences were evident in place of residence. A significant association was identified only in Papua, where children living in urban areas had lower odds of being stunted at birth than those residing in rural areas (cOR: 0.47; 95% CI: 0.24–0.91). Household economic status also showed regional variation. In Sumatra, children from households in the lowest wealth quintile had 1.59 times higher odds of stunting at birth than those from households in the highest quintile (cOR: 1.59; 95% CI: 1.00–2.52). Conversely, in Kalimantan, children from households in the upper-middle wealth quintile exhibited higher odds of stunting at birth compared with those from the highest wealth quintile (cOR: 1.94; 95% CI: 1.15–3.29). Parental education was generally not associated with stunting at birth across regions. Nevertheless, a significant association was observed in Maluku, where children whose fathers had secondary education had lower odds of being stunted at birth than those whose fathers had higher education (cOR: 0.26; 95% CI: 0.09–0.69). Maternal nutritional knowledge likewise demonstrated regional variation. In Java–Bali, low maternal nutritional knowledge was associated with increased odds of stunting at birth (cOR: 1.65; 95% CI: 1.03–2.63), whereas an inverse association was identified in Maluku (cOR: 0.20; 95% CI: 0.07–0.53).

**Table 4.** Bivariate Analysis of Sociodemographic, Household, and Environmental Determinants of Stunting at Birth Across Regions in Indonesia

| Variabel | Sumatera | Jawa-Bali | Nusa Tenggara | Kalimantan | Sulawesi | Maluku | Papua |
| --- | --- | --- | --- | --- | --- | --- | --- |
|  | cOR (95 % CI) | cOR (95 % CI) | cOR (95 % CI) | cOR (95 % CI) | cOR (95 % CI) | cOR (95 % CI) | cOR (95 % CI) |
| **Place of residence** |  |  |  |  |  |  |  |
| Urban | 0,82 (0,62-1,09) | 0,82 (0,62-1,08) | 0,98 (0,59-1,62) | 1,16 (0,80-1,67) | 1,01 (0,75-1,36) | 0,87 (0,38-2,00) | 0,47 (0,24-0,91)* |
| Rural | Ref | Ref | Ref | Ref | Ref | Ref | Ref |
| **Household economic status** |  |  |  |  |  |  |  |
| Lowest Quintile | 1,59 (1,00-2,52)* | 0,96 (0,64-1,44) | 2,12 (0,59-7,56) | 0,93 (0,50-1,71) | 1,38 (0,87-2,18) | 3,36 (0,66-16,96) | 0,99 (0,39-2,53) |
| Lower-Middle Quintile | 1,33 (0,83-2,12) | 0,97 (0,68-1,38) | 1,08 (0,27-4,31) | 1,31 (0,73-2,35) | 0,84 (0,51-1,38) | 1,55 (0,25-9,28) | 1,95 (0,64-5,93) |
| Middle Quintile | 1,19 (0,76-1,84) | 1,06 (0,72-1,55) | 1,25 (0,30-5,10) | 1,04 (0,60-1,81) | 0,81 (0,50-1,31) | 3,58 (0,68-18,85) | 0,99 (0,31-3,12) |
| Upper Middle Quintile | 1,09 (0,68-1,77) | 1,06 (0,70-1,62) | 2,07 (0,49-8,60) | 1,94 (1,15-3,29)* | 0,86 (0,49-1,52) | 3,04 (0,52-17,67) | 1,40 (0,52-3,80) |
| Highest Quintile | Ref | Ref | Ref | Ref | Ref | Ref | Ref |
| **Maternal education** |  |  |  |  |  |  |  |
| Primary education | 1,17 (0,81-1,69) | 0,88 (0,62-1,25) | 1,35 (0,68-2,66) | 0,97 (0,55-1,68) | 1,09 (0,72-1,67) | 1,53 (0,44-5,31) | 0,94 (0,38-2,30) |
| Secondary education | 0,95 (0,65-1,39) | 0,94 (0,66-1,35) | 1,57 (0,76-3,25) | 0,85 (0,47-1,54) | 1,02 (0,67-1,54) | 1,17 (0,46-2,92) | 0,91 (0,39-2,10) |
| Higher education | Ref | Ref | Ref | Ref | Ref | Ref | Ref |
| **Paternal education** |  |  |  |  |  |  |  |
| Primary education | 0,88 (0,57-1,36) | 0,89 (0,60-1,32) | 1,27 (0,56-2,86) | 1,26 (0,65-2,44) | 0,96 (0,61-1,52) | 1,25 (0,47-3,36) | 0,90 (0,33-2,38) |
| Secondary education | 0,84 (0,54-1,30) | 0,88 (0,60-1,29) | 1,47 (0,63-3,40) | 0,83 (0,43-1,60) | 1,06 (0,67-1,70) | 0,26 (0,09-0,69)** | 0,94 (0,40-2,23) |
| Higher education | Ref | Ref | Ref | Ref | Ref | Ref | Ref |
|  | cOR (95% CI) | cOR (95% CI) | cOR (95% CI) | cOR (95% CI) | cOR (95% CI) | cOR (95% CI) | cOR (95% CI) |
| **Maternal knowledge** |  |  |  |  |  |  |  |
| Low | 0,93 (0,58-1,50) | 1,65 (1,03-2,63)* | 1,17 (0,61-2,24) | 0,77 (0,45-1,32) | 0,99 (0,63-1,55) | 0,20 (0,07-0,53)** | 1,38 (0,36-5,19) |
| High | Ref | Ref | Ref | Ref | Ref | Ref | Ref |
| **Household crowding** |  |  |  |  |  |  |  |
| Does not meet standard | 1,31 (0,93-1,83) | 1,31 (0,95-1,80) | 1,60 (0,94-2,72) | 0,87 (0,53-1,41) | 1,14 (0,82-1,57) | 1,71 (0,47-6,14) | 1,47 (0,71-3,05) |
| Meets standard | Ref | Ref | Ref | Ref | Ref | Ref | Ref |
| **Access to drinking water** |  |  |  |  |  |  |  |
| Surface water | 0,87 (0,19-3,84) | Empty | 4,30 (0,48-38,38) | 0,94 (0,30-2,93) | Empty | Empty | 0,36 (0,04-3,25) |
| Unimproved | 0,98 (0,50-1,94) | 0,95 (0,41-2,22) | 0,68 (0,21-2,15) | 0,88 (0,22-3,47) | 0,42 (0,16-1,08) | Empty | 0,74 (0,08-6,36) |
| Limited | 0,85 (0,36-2,01) | 0,65 (0,32-1,35) | 2,59 (1,25-5,39)* | 0,77 (0,33-1,79) | 1,14 (0,61-2,15) | 1,75 (0,52-5,84) | 0,34 (0,87-1,36) |
| Basic | 0,91 (0,68-1,20) | 1,10 (0,84-1,45) | 0,45 (0,26-0,79)** | 0,92 (0,61-1,39) | 1,14 (0,79-1,65) | 0,86 (0,33-2,19) | 1,15 (0,57-2,34) |
| Safety managed | Ref | Ref | Ref | Ref | Ref | Ref | Ref |
| **Access to sanitation** |  |  |  |  |  |  |  |
| Open defecation | 0,69 (0,14-3,36) | 0,09 (0,01-0,48)** | Empty | Empty | 1,84 (0,67-5,06) | Empty | Empty |
| Unimproved | 0,60 (0,12-2,82) | 0,73 (0,31-1,72) | Empty | 1,16 (0,55-2,42) | 0,54 (0,11-2,62) | Empty | 1,83 (0,22-14,88) |
| Limited | 0,53 (0,09-2,90) | 0,58 (0,23-1,46) | 2,78 (1,55-4,98)* | 1,20 (0,54-2,66) | 1,17 (0,72-1,88) | 0,15 (0,01-1,21) | 1,00 (0,24-4,06) |
| Basic shared | 0,49 (0,09-2,44) | 0,58 (0,24-1,43) | 1,54 (0,56-4,19) | 0,56 (0,17-1,86) | 1,23 (0,70-2,15) | 0,85 (0,23-3,06) | 0,60 (0,15-2,31) |
| Basic private | 0,53 (0,12-2,29) | 0,58 (0,27-1,25) | Omitted | Omitted | Omitted | Omitted | Omitted |
| Safely managed | Ref | Ref | Ref | Ref | Ref | Ref | Ref |

**Table 5.**
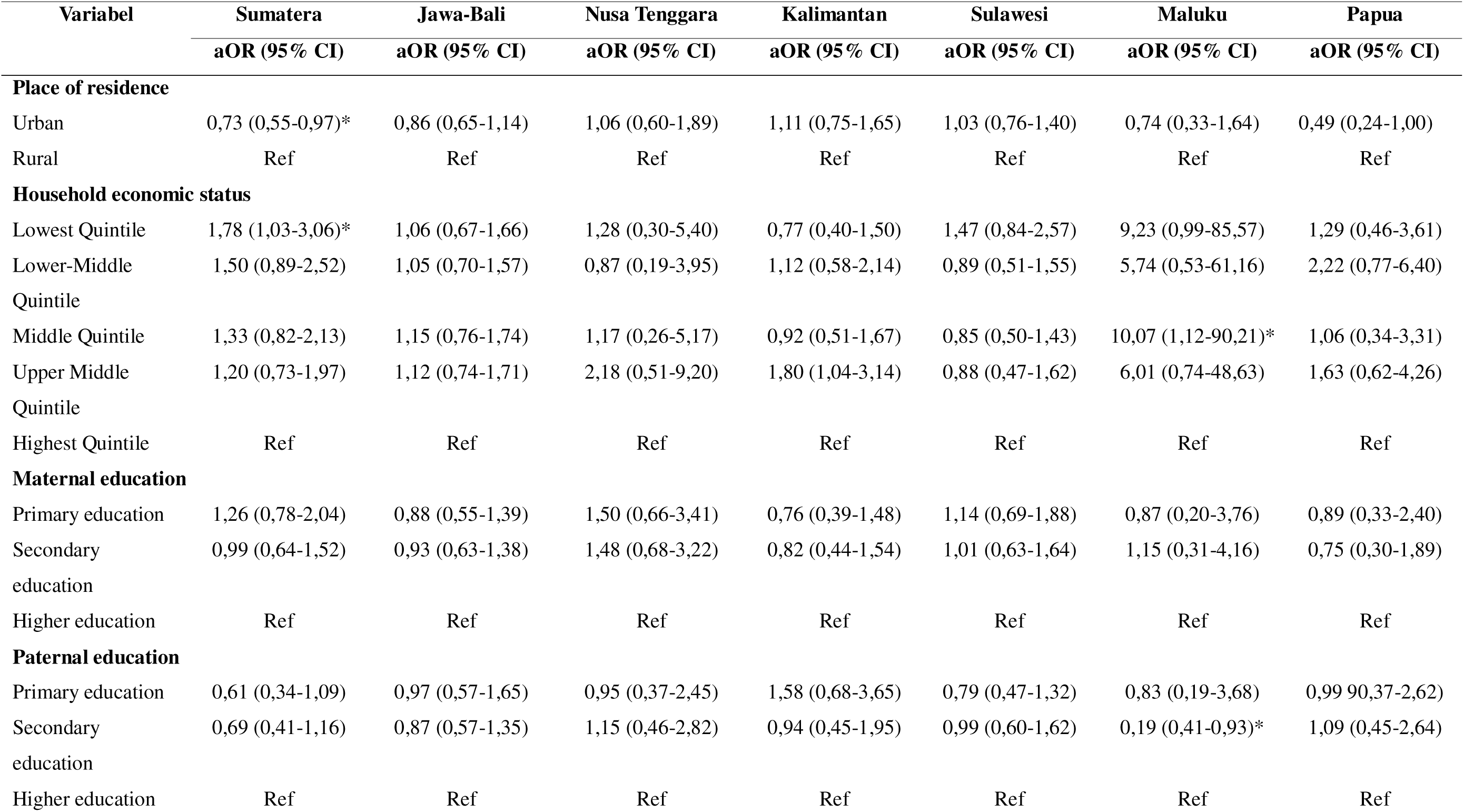
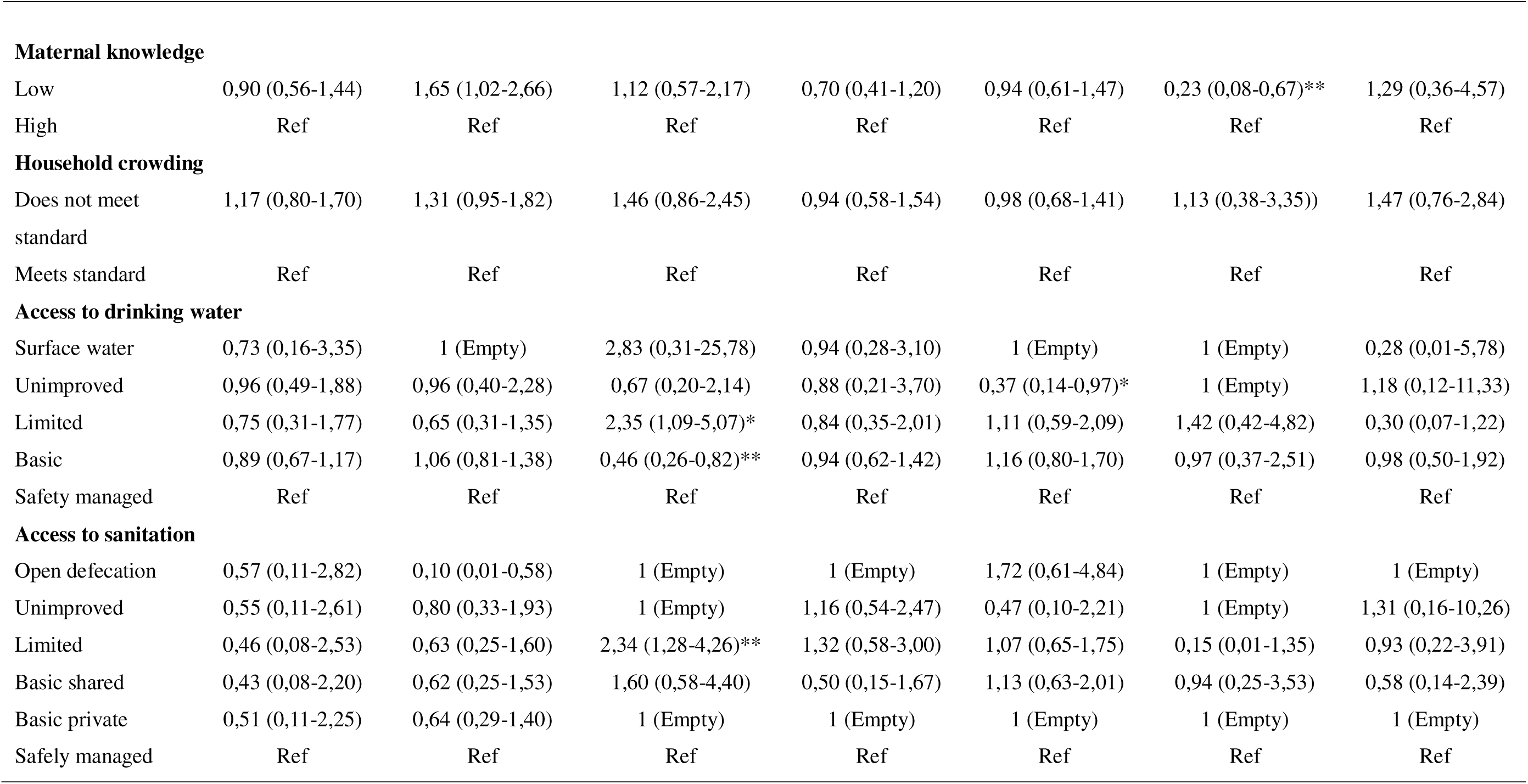
Multivariable Analysis of Socioeconomic, Household, and Environmental Determinants of Stunting at Birth Across Regions in Indonesia

The adjusted associations between sociodemographic, household, and environmental factors and stunting at birth across seven regions of Indonesia are presented in Table 4. Compared with the bivariate analysis, several associations remained statistically significant after adjustment, whereas others were attenuated and no longer reached statistical significance, suggesting the influence of potential confounding factors. Regional differences were evident in place of residence. After adjustment, urban residence was associated with lower odds of stunting at birth in Sumatra compared with rural residence (aOR: 0.73; 95% CI: 0.55–0.97). This association emerged only in the multivariable analysis, while the significant association previously observed in Papua in the bivariate analysis was no longer evident after adjustment. Household economic status also demonstrated regional variation. In Sumatra, children from households in the lowest wealth quintile had 1.78 times higher odds of stunting at birth than those from households in the highest quintile (aOR: 1.78; 95% CI: 1.03–3.06). In contrast, the association identified among children in the upper-middle wealth quintile in Kalimantan was no longer statistically significant after adjustment.

Parental education was generally not associated with stunting at birth across regions. Nevertheless, a significant association persisted in Maluku, where children whose fathers had secondary education exhibited lower odds of stunting at birth than those whose fathers had higher education (aOR: 0.19; 95% CI: 0.04–0.93). Maternal nutritional knowledge likewise showed consistent regional patterns. Low maternal nutritional knowledge remained associated with increased odds of stunting at birth in Java–Bali (aOR: 1.65; 95% CI: 1.02–2.66), whereas the inverse association observed in Maluku persisted after adjustment (aOR: 0.23; 95% CI: 0.08–0.67). Environmental factors remained particularly important in Nusa Tenggara. Significant associations identified in the bivariate analysis persisted in the adjusted model, with households using limited drinking water services exhibiting more than twice the odds of stunting at birth compared with those using safely managed drinking water services (aOR: 2.35; 95% CI: 1.09–5.07). Conversely, access to basic drinking water services was associated with lower odds of stunting at birth (aOR: 0.46; 95% CI: 0.26–0.82). Similarly, households with limited sanitation services had higher odds of stunting at birth than those with safely managed sanitation (aOR: 2.34; 95% CI: 1.28–4.26). By contrast, the significant association between open defecation and stunting at birth observed in Java–Bali in the bivariate analysis was no longer apparent after adjustment. No significant associations were identified for household crowding in any region in either the bivariate or multivariable analyses.

## Discussion

This study examined the determinants of stunting at birth at both the national and regional levels in Indonesia. At the national level, household crowding was the only factor that remained significantly associated with stunting at birth after adjustment. However, the regional analysis revealed substantial heterogeneity in associated factors. Urban residence and household economic status were associated with stunting at birth in Sumatra, low maternal nutritional knowledge emerged as an important determinant in Java–Bali, and environmental factors, particularly drinking water and sanitation services, were strongly associated with stunting at birth in Nusa Tenggara. These findings suggest that although some determinants may be identified at the national level, their effects are not uniformly distributed across regions. Consequently, national estimates may mask important local patterns that are relevant for the design of targeted stunting prevention strategies.

Household crowding was the only factor that remained significantly associated with stunting at birth at the national level. This finding suggests that household living conditions may influence fetal growth and development even before birth. Overcrowded households are often characterized by limited living space, reduced environmental quality, and increased exposure to infectious diseases, all of which may adversely affect maternal health during pregnancy. In addition, household crowding may reflect broader socioeconomic disadvantage and poor housing conditions. According to the World Health Organization, overcrowded housing can negatively affect health through increased transmission of infectious diseases, reduced ventilation, and compromised living conditions [12]. Similar findings have been reported in Indonesia, where household crowding was significantly associated with stunting among children [13]. Interestingly, this association was not observed in any individual region after stratified analysis. This discrepancy may indicate that the effect of household crowding is distributed across multiple regions and becomes apparent only when data are analyzed at the national level. Alternatively, the loss of statistical significance may reflect reduced statistical power following regional stratification. These findings suggest that household crowding may operate as a structural determinant of stunting at birth across Indonesia rather than as a region-specific risk factor.

The substantial regional variation observed in this study is consistent with Indonesia’s diverse geographical, socioeconomic, and infrastructural conditions. Marked differences exist across regions in terms of poverty, food security, maternal healthcare access, educational attainment, and environmental infrastructure. These disparities may influence maternal nutrition and fetal growth through different pathways, resulting in distinct determinants of stunting at birth across regions. Therefore, understanding regional variation is essential because interventions based solely on national averages may fail to address local risk factors operating within specific settings.

In Sumatra, urban residence was associated with lower odds of stunting at birth compared with rural residence. This finding may reflect differences in access to maternal healthcare services, transportation infrastructure, nutrition programs, and health information between urban and rural communities. Geographic barriers and limited healthcare availability in rural areas may reduce access to antenatal care and maternal nutrition services, thereby increasing the risk of impaired fetal growth. However, evidence regarding the relationship between place of residence and stunting remains inconsistent. A previous study by Gonete et al. reported no significant association between rural residence and stunting after adjustment for other determinants. These differences suggest that the influence of place of residence may depend on local socioeconomic and environmental conditions rather than the urban–rural classification alone [6]. In Indonesia, urban advantages may also vary considerably across regions because access to healthcare and public infrastructure is not evenly distributed.

Household economic status also remained an important determinant in Sumatra, where children from households in the lowest wealth quintile had significantly higher odds of stunting at birth than those from the highest wealth quintile. Household economic resources influence maternal and child health through multiple pathways, including food security, dietary diversity, healthcare utilization, and living conditions. Women from poorer households may face greater barriers in obtaining adequate nutrition during pregnancy, increasing the likelihood of fetal growth restriction. This finding is consistent with previous evidence from Indonesia showing that children from low-income households are more likely to experience stunting than those from wealthier households [14]. Similarly, Beal et al. reported that household socioeconomic conditions remain a key determinant of child growth and nutritional status in Indonesia [15].

Low maternal nutritional knowledge was associated with increased odds of stunting at birth in Java–Bali. Maternal knowledge plays a critical role in shaping health-related decision-making during pregnancy, including dietary practices, healthcare-seeking behavior, utilization of antenatal care services, and adherence to nutritional recommendations. Mothers with better nutritional knowledge are generally more likely to adopt behaviors that support adequate fetal growth and development. Previous studies have shown that limited understanding of nutrition and child growth may reduce the adoption of preventive health behaviors. Research conducted in Indonesia found that some mothers perceive stunting primarily as a hereditary condition rather than a preventable health problem, potentially reducing motivation to engage in preventive practices [16]. In addition, improved maternal knowledge has been associated with greater utilization of maternal and child health services and healthier caregiving behaviors, which may contribute to improved birth outcomes and child growth. Interestingly, an inverse association was observed in Maluku, where low maternal nutritional knowledge appeared to be associated with lower odds of stunting at birth. Given the relatively small sample size and wide confidence intervals observed in this region, this finding should be interpreted cautiously and may reflect residual confounding or statistical instability rather than a true protective effect.

Environmental factors emerged as important determinants of stunting at birth in Nusa Tenggara. Households with limited drinking water services had more than twice the odds of stunting at birth compared with those having safely managed drinking water, while access to basic drinking water services was associated with lower odds of stunting at birth. Similarly, limited sanitation services remained significantly associated with increased odds of stunting at birth after adjustment. These findings suggest that inadequate access to essential water, sanitation, and hygiene (WASH) services may contribute to growth faltering beginning in the prenatal period.

Several biological mechanisms may explain these associations. Access to safe drinking water and adequate sanitation plays an important role in maintaining maternal health during pregnancy and preventing infections that may adversely affect fetal growth. Inadequate WASH services increase exposure to gastrointestinal and other environmentally related infections, which can impair nutrient absorption, increase nutritional requirements, and worsen maternal nutritional status. Previous studies have shown that households lacking adequate drinking water and sanitation facilities face a greater risk of stunting than those with access to safe and improved services [17]. Furthermore, poor sanitation has been linked to maternal health complications during pregnancy, including conditions associated with infection and adverse pregnancy outcomes that may affect fetal growth [18]. Recurrent infections and chronic inflammation may contribute to placental dysfunction and fetal growth restriction, thereby increasing the risk of stunting at birth.

The strong associations observed in Nusa Tenggara may reflect persistent disparities in access to basic environmental infrastructure across Indonesian regions. Descriptive findings from this study showed that Nusa Tenggara had one of the highest proportions of households relying on limited drinking water and sanitation services. These conditions may increase vulnerability to waterborne and sanitation-related diseases, making environmental factors more influential in this region than in other parts of Indonesia. Similar findings have been reported in Ethiopia, where the use of unimproved sanitation facilities was associated with a twofold increase in the odds of stunting among children [19]. While drinking water and sanitation were not significant determinants at the national level, the regional findings suggest that their effects may be context-specific and become more apparent in areas where access to basic services remains limited. Therefore, efforts to reduce stunting at birth in Nusa Tenggara should be complemented by improvements in safe drinking water and sanitation infrastructure alongside maternal nutrition interventions.

This study has several limitations. First, the cross-sectional design precludes causal inference between sociodemographic, household environmental factors, and stunting at birth. Second, some regional analyses, particularly in Maluku and Papua, were based on relatively small sample sizes, resulting in wide confidence intervals and reduced statistical precision. Nevertheless, the study utilized nationally representative data from the 2024 Indonesian Nutritional Status Survey and applied survey-weighted analyses, providing robust evidence on regional disparities in determinants of stunting at birth across Indonesia.

## Conclussion

This study identified substantial regional variation in the determinants of stunting at birth across Indonesia. At the national level, household crowding was the only factor significantly associated with stunting at birth after adjustment. However, regional analyses revealed distinct patterns, with socioeconomic factors and place of residence emerging as important determinants in Sumatra, maternal nutritional knowledge in Java–Bali, and drinking water and sanitation services in Nusa Tenggara. These findings suggest that the determinants of stunting at birth are context-specific and may not be adequately captured by national-level estimates alone. Therefore, strategies to prevent stunting beginning in the prenatal period should incorporate regional characteristics and prioritize interventions according to local needs and risk factors.

## Data Availability

All data produced in the present study are available upon reasonable request to the authors

## Author Contributions

Fawwiz Aulya Amin: Conceptualization, Methodology, Formal analysis, Data curation, Investigation, Visualization, Writing – original draft, Writing – review & editing. Siti Helmyati: Supervision, Validation, Writing – review & editing. Vicka Oktaria: Supervision, Validation, Writing – review & editing.

## Acknowledgments

The authors would like to thank the Ministry of Health of the Republic of Indonesia for providing access to the 2024 SSGI dataset. The authors also acknowledge Universitas Gadjah Mada for academic support during the conduct of this study.

## Competing Interests

The authors declare that they have no competing interests.

The word count exceeds the recommended limit due to the inclusion of stratified analyses across seven regions of Indonesia. These analyses are central to the study objectives and required additional reporting of region-specific results and interpretations. We have minimized unnecessary text and retained only information essential for understanding the findings.

